# MAPEVA-ScR: a four-phase framework integrating transparent rule-based text mining, evidence gap mapping and a mandatory human validation gate for scoping reviews — framework development and empirical validation in occupational health

**DOI:** 10.64898/2026.09.08.26362521

**Authors:** César Jesús Eras Lévano

## Abstract

**Background:** Automation is increasingly used to reduce the screening workload of evidence syntheses, but its error is real, variable and rarely reported. The 2025 joint position statement of Cochrane, Campbell, JBI and the Collaboration for Environmental Evidence mandates human oversight of artificial intelligence in evidence synthesis, yet — as of today — no international guideline requires quantitative measurement and reporting of automated-screening error against a human gold standard (1).

**Methods:** We developed MAPEVA-ScR (MAPeo de EVidencia con VAlidación humana), a four-phase framework for scoping reviews: (A) datalake construction with deterministic, traceable deduplication; (B) transparent, record-level auditable rule-based text mining with confidence tiers; (C) evidence gap mapping that feeds back human screening prioritisation; and (D) a mandatory human validation gate verifying 100% of the candidate corpus and issuing a quantitative screening error report (per-tier precision, confusion matrix, false-positive families, documented changes to conclusions). The framework was empirically validated on a scoping review of epilepsy and occupational fitness (2015–2025; PubMed, Scopus, Web of Science): a funnel of 2,008 → 1,435 → 1,114 → 230 records was reduced to 127 human-confirmed inclusions (present study).

**Results:** The rule-based filter achieved a global precision of 55.2% (Tier A 75.5%, Tier B 37.5%, Tier C 45.8%); human validation identified 103 false positives classifiable into seven reproducible error families, and 107 of 127 included records required human correction of automated thematic labels. Without the gate, the review’s central thematic conclusion would have been published reversed: automated labelling ranked Aptitude/employability third (12.6% of records), whereas after validation it ranked first (73.2%) (present study).

**Conclusions:** A mandatory human validation gate with quantitative error metrics closes a reporting gap that current guidelines leave open: in the present study it prevented publication of an inverted synthesis. We propose the screening error report as a minimum reporting standard for any evidence synthesis using automated screening.

**Registration and protocol:** Not applicable (methods development and validation study reporting original empirical data; no health intervention outcomes).

**AI use disclosure:** Generative AI assisted with language editing under full human verification (2); the MAPEVA-ScR pipeline uses deterministic rule-based text mining, with no generative AI in screening decisions.

## 1. Introduction

### 1.1 The screening bottleneck and the automation turn

Screening of titles and abstracts is the most labour-intensive stage of systematic and scoping reviews. Semi-automation promises relief: a systematic review of text-mining approaches estimated workload reductions of 30–70%, “sometimes at the cost of up to a 5% loss of relevant studies” (3). The Cochrane Handbook reiterates that machine learning can reduce the screening burden by at least 30%, and possibly by more than 90%, while warning that automatic exclusion of records via active learning “has not been recommended for use in Cochrane Reviews” (4). The efficiency promise is real, but conditional on an error rate that is rarely quantified locally.

That error is neither hypothetical nor constant: a sensitive GPT-3.5 rule reached 94.6–99.8% sensitivity with specificity as low as 2.2% (5); the Cochrane Rapid Reviews Methods Group reports incorrect AI inclusion decisions ranging from 0 to 29% (median 10%) (2). Nor is error exclusively machine-made: single-reviewer abstract screening misses approximately 13% of relevant studies (6), and machine learning recovered 33–90% of records that a second human reviewer had missed, indicating genuine but heterogeneous human–machine complementarity (7).

Large language models (LLMs) introduce failure modes of their own: bibliographic hallucination rates of 39.6% (GPT-3.5), 28.6% (GPT-4) and 91.4% (Bard) (8); fabricated citations in 55% and 18% of generated reference lists depending on model version (9); and a field verdict of “on the rise, but not yet ready for use” (10). Automation bias — the tendency of human reviewers to over-trust automated outputs — is well recognised in the human-factors literature but remains barely operationalised in screening practice.

### 1.2 The governance response — and its limit

The evidence synthesis community has responded with governance instruments. In 2025, Cochrane, the Campbell Collaboration, JBI and the Collaboration for Environmental Evidence issued a joint position statement on artificial intelligence (AI) use in evidence synthesis, mandating human oversight, transparent reporting of tool name, version and purpose, justification of tool choice with evidence of methodological soundness, and a description of how the tool was validated or piloted (1). The RAISE (Responsible use of AI in evidence SynthEsis) framework adds role-specific guidance: RAISE 1 on roles and responsibilities, RAISE 2 on tool evaluation with performance metrics, and RAISE 3 on tool selection for a specific synthesis (11).

We do not claim that human oversight is our invention; the 2025 joint statement already requires it (1). Our argument concerns what oversight is not yet required to produce. As of today, no current guideline mandates measuring and quantitatively reporting the error of the automated screening component against a human gold standard: no required confusion matrix, no per-stratum precision, and no obligation to document which conclusions changed after human validation. PRISMA 2020 and its explanation and elaboration document ask — only for internally derived machine-learning classifiers — that authors “describe” the validation performed, narratively, without metrics or acceptance thresholds (12) (13). This is a gap as of today, not an eternal void: PRISMA-trAIce has been proposed as a dedicated checklist (14) and RAISE 2 already lists performance metrics as good practice (11). Until such requirements are binding, however, a team may comply fully with existing guidance while publishing screening results whose error was never measured.

### 1.3 Contribution statement

This paper develops and empirically validates MAPEVA-ScR (MAPeo de EVidencia con VAlidación humana applied to scoping reviews) and makes three contributions. First, an architecture: to our knowledge — based on a broad but not formally systematic positioning search, to be verified in Scopus and Web of Science before submission — no published framework combines a datalake construction phase, transparent record-level auditable rule-based tier stratification, evidence gap mapping that feeds back human screening prioritisation, and a mandatory human validation gate, in a pipeline designed for non-programmer occupational health researchers and implemented in Python/pandas with auditable rules.

Second, a reporting standard: a mandatory “screening error report” comprising per-tier precision, a confusion matrix of automated tier against human judgement, a taxonomy of false-positive families, and a log of human corrections — none of which any current guideline requires. Third, empirical evidence of the risk: to our knowledge the first quantified documentation of a reversed thematic conclusion caused by unvalidated screening in a scoping review (present study).

The framework is positioned against its closest precedents. Pham et al. published a text-mining workflow with performance metrics (sensitivity 88–89%, precision 71–72%, F1 79%) (15); we extend that precedent with the datalake, the feedback gap mapping, the mandatory human gate, and an occupational health application. SWIFT-Review and SWIFT-Active Screener combine explicit rule filters with mapping in environmental health (16) (17); we add the validation gate with reported metrics and the reporting standard, in the JBI/PRISMA-ScR scoping context. Active-learning tools such as ASReview and RobotAnalyst (18) (19) prioritise records through grey-box classifiers; our rules are auditable record by record, and the gate can follow such prioritisation — we complement rather than compete. The joint statement and RAISE set normative principles (1) (11); MAPEVA-ScR delivers an operational pipeline with quantitative metrics. Finally, unlike EPPI-Mapper-style gap maps (20), our map feeds back into screening: it decides what the human reads first.

The stakes are concrete. In our validation case (a scoping review of epilepsy and occupational fitness, 2015–2025; funnel 2,008 → 1,435 → 1,114 → 230 → 127 records), the rule-based filter achieved a global precision of 55.2%, and automated thematic labelling would have ranked Aptitude/employability third (12.6%) when human validation placed it first (73.2%): the thematic conclusion would have been published reversed (present study). Yet a team reporting that pipeline could have complied nominally with PRISMA 2020, PRISMA-ScR and the 2025 joint statement, because none requires the error to be measured. The remainder of the article is organised as follows: Section 2 reviews screening tools, gap mapping and guidelines; Section 3 specifies the four phases of MAPEVA-ScR; Section 4 reports the empirical validation; Section 5 discusses findings, limitations and implications; Section 6 concludes.

## 2. Background and Related Work

### 2.1 Automated and semi-automated screening tools

#### 2.1.1 Classifiers and active learning

The dominant paradigm is the machine learning (ML) classifier with active learning: a human labels records and the model continuously re-ranks the corpus. ASReview, the most visible open-source implementation, reported mean work saved over sampling at 95% recall (WSS@95) of ≈83% (range 67–92%), finding 95% of relevant records after screening 8–33% of the corpus; these figures derive, however, from retrospective simulation on 15 datasets, not from prospective evaluation against a blinded gold standard (18). Abstrackr, a support vector machine classifier with active learning, achieved sensitivity ≥0.75 with widely varying specificity (0.19–0.90) and a median workload saving of 67% (21); its first prospective evaluation estimated savings of 40–57% (22). RobotAnalyst sustained 95% recall while reducing the corpus in 19 of 22 collections in the first large-scale user study (43,610 decisions) (19), and RobotReviewer extended ML to PICO extraction and risk-of-bias assessment with domain-variable performance (23). Two limitations recur across this trajectory: the decision logic is a grey- or black-box model not auditable record by record, and validation remains a retrospective research exercise rather than a mandatory gate within the review itself.

#### 2.1.2 Rule-adjacent and plaVorm approaches

SWIFT-Review, developed for environmental health at NIEHS/EPA, applies explicit, user-inspectable rule-based topic filters with mapping visualisations (16); its successor, SWIFT-Active Screener, adds active learning with integrated recall estimation (17). These are the closest precedents in spirit, yet neither imposes a mandatory human validation gate with reported error metrics, nor have they been applied to scoping reviews (ScRs) under JBI/PRISMA-ScR methodology. EPPI-Reviewer offers ML priority screening and EPPI-Mapper produces gap maps, but validation remains optional rather than gated; Khalil et al.’s scoping review confirms that most platforms lack published validation of their screening components (20), and the AHRQ-commissioned evaluation found tool performance contingent on review characteristics (24). Rayyan, widely used in ScRs, was published descriptively, without formal validation of its ML ranking (25). Cochrane’s Screen4Me combines crowdsourcing, an RCT classifier and expert verification — accurate for identifying randomised trials, not for generic eligibility screening (26) (27).

#### 2.1.3 Text-mining workflow with metrics — the nearest methodological precedent

Pham et al. published in this journal a semi-automated text-mining workflow reporting sensitivity 88–89%, precision 71–72% and F1 79% across one systematic review (SR) and one ScR (15). This is, to our reading, the nearest methodological precedent: it shows that transparent text mining with reported metrics is publishable, but it addresses screening only

— no datalake, no evidence gap mapping, no mandatory human gate — and is the precedent MAPEVA-ScR most directly extends.

#### 2.1.4 LLMs for screening (2023–2026)

Since 2023, large language models (LLMs) have been evaluated for title/abstract screening with strikingly heterogeneous results, varying sharply by model and prompt (28): sensitivity 0.95 with specificity 0.65 for ChatGPT (29); accuracy ≈0.91 with recall 0.76–0.91 for GPT-4 (30); GPT-4 accuracy ≥90%, with LLMs outperforming ASReview and Abstrackr on sensitivity and AUC (31). Independent analyses qualify these headline numbers: GPT-4 reaches human-level performance only at full text, title/abstract screening being degraded by class imbalance and chance agreement (32), and a benchmark of 18 LLMs across three reviews found modest mean Matthews correlation coefficients (≈0.31–0.35) with a criteria×model interaction (33); a 2025 scoping review concludes the field is “on the rise, but not yet ready for use” (10). The synthesis is consistent: screening error is real, variable and unreported by default — no mainstream tool emits a per-review confusion matrix against a human gold standard.

### 2.2 Evidence gap mapping

Evidence gap maps (EGMs), formalised in the 3ie/Campbell methodology (Snilstveit et al. 2016) (34), display the distribution of evidence across an intervention–outcome framework to guide research prioritisation. The methodology has matured unevenly: a 2024 review of Campbell EGMs found that only 33% of maps methodise gap identification (35) — an inconsistency that itself argues for a formal, auditable mapping phase. Living EGMs extend the format to continuously updated evidence bases (36), and Big Picture Reviews propose broad mapping as a first-pass synthesis product (37). For the present argument, the decisive limitation is structural rather than technical: mapping is decoupled from screening. Maps are produced after screening, so they neither feed back into human prioritisation nor quantify the error accumulated upstream; the map simply inherits whatever the filter removed or contaminated. A mapping phase that precedes final human validation, and is itself corrected by it, is absent from the EGM literature we reviewed.

### 2.3 Guidelines and the regulatory gap

#### 2.3.1 What each guideline requires

PRISMA 2020 (items 8, 9, 11, 16a) requires reporting details of automation tools used, and its flow diagram separately counts “records marked as ineligible by automation tools” (12). Its explanation and elaboration document goes furthest, asking — but only for internally derived ML classifiers — that authors “describe what internal or external validation was done”: a narrative request, with no required metrics or thresholds (13). PRISMA-S governs search reporting, not screening (38); PRISMA-ScR, predating PRISMA 2020, never mentions automation (39). The JBI Manual mandates two or more independent reviewers, a pilot of eligibility criteria and third-reviewer arbitration — a 100%-human paradigm whose only contemplated validation is of criteria between humans, not algorithm performance (40) (41). The Cochrane Handbook (§4.6.6.2) accepts up to ∼5% sensitivity loss as a known cost without requiring local quantification, and warns that automatic exclusion via active learning “has not been recommended for use in Cochrane Reviews” (4). MECIR standard C39 — the ecosystem’s only explicitly mandatory tier — presupposes human double screening; no MECIR standard addresses artificial intelligence (AI) (42), and the updated Campbell reporting standard mentions validation (“e.g., 10% review by human”) only as a conditional example (43). The 2025 joint position statement of Cochrane, Campbell, JBI and CEE mandates human oversight and transparent reporting of tool, version, purpose and validation approach, but entirely in “should/report/describe” language (1); RAISE lists performance metrics as good practice for tool developers and evaluators (RAISE 2), not as a binding requirement for individual syntheses (11). The Cochrane Rapid Reviews Methods Group prohibits full automation in rapid reviews, framing AI as quality assurance, and introduces an “AI Use Disclosure” section (2); NICE holds that AI must augment, not replace, human involvement (44).

#### 2.3.2 The demonstrated gap

As of today, no current international guideline or standard requires reporting the precision or sensitivity of automated screening validated against a human gold standard with defined quantitative metrics — confusion matrix, false-positive/false-negative rates, minimum acceptance thresholds. Practice evidence corroborates the consequence: across 2,271 Cochrane, Campbell and CEE syntheses (2017–2024), only ∼5% report explicit ML use, ∼90% of ML-tool users do not clarify whether the ML function was used, and standards give “minimal” coverage to automation reporting (43). Two nuances are required. First, human oversight is already mandatory under the 2025 joint statement (1); what is missing as of today is the obligation to measure and quantitatively report the automated component’s error — per-tier precision, confusion matrix, documentation of which conclusions changed after validation. Second, the gap is closing: RAISE is in active development (11), the PRISMA-trAIce checklist has been proposed for transparent AI reporting (14), and AI Use Disclosure is already required in rapid reviews (2). The gap is therefore the absence of a mandatory, quantitative requirement as of today, not an absence of recommendations.

#### 2.3.3 Occupational health as application domain

In occupational health, AI appears as the object of scoping reviews — syntheses of AI for workforce health and work disability prevention (45) and of AI in occupational rehabilitation (46) — not as a validated screening method. The nearest methodological work, a systematic review of natural language processing applied to occupational injury narratives, mines accident reports as data rather than conducting the review itself (47). We found no validated methodological application of automated screening in the domain.

### 2.4 Positioning summary

To our knowledge, no published framework combines (a) transparent, record-level auditable rule-based tier stratification, (b) evidence gap mapping that feeds back human screening prioritisation, and (c) a mandatory human validation gate with reported error metrics, applied to scoping reviews; our positioning search was broad — spanning the tools, guidelines and frameworks cited above — but was not a formal systematic search, and will be verified in Scopus and Web of Science before submission. Adjacent frameworks confirm the niche by contrast: ScoRBA structures ScRs bibliometrically, without rule-based mining, gap mapping or a validation gate (48), and the rapid reviews methods series recommends semi-automation with sampling-based human verification — sound guidance, but not a mandatory gate with quantified filter error (49). Section 3 specifies the framework that occupies this niche.

## 3. The MAPEVA-ScR Framework

### 3.1 Design principles and overview

MAPEVA-ScR (MAPeo de EVidencia con VAlidación humana — evidence mapping with human validation — for scoping reviews) is a four-phase framework designed for a specific user: the occupational or public health researcher who is not a programmer. The reference implementation is pure Python/pandas; every automated decision is an explicit, versioned rule explainable record-by-record to a second human reviewer. Four design principles govern the framework. **P1 — Transparency over black-box performance**: we chose explicit, auditable rules over grey-box active learning (18), accepting lower nominal automation gains in exchange for record-level explainability. **P2 — Human sight of the entire candidate corpus**: the human verifier sees the whole candidate corpus and no candidate record is machine-excluded without human sight. This principle applies to the post-filter candidate corpus: the Phase B filter prioritises rather than definitively excludes — non-retained records remain in the datalake, inspectable on demand — and the unmeasured sensitivity of that upstream filter is declared as a limitation (§5.3). The principle stands in contrast to the ∼5% sensitivity loss the Cochrane Handbook tolerates, and respects its caution against automatic exclusion via active learning (4). **P3 — The map prioritises the human, not vice versa**: the evidence gap map feeds back into screening, deciding what the human reads first. **P4 — Validation is a gate, not an option**: no output of Phases A–C may be used analytically until the Phase D gate has been passed and its error report produced. No acronym collisions with MAPEVA-ScR were detected in a broad positioning search; a formal Scopus check will precede submission.

Figure 1 summarises the framework. Phase A builds a deduplicated bibliographic datalake; Phase B applies rule-based text mining assigning each record a confidence tier and provisional thematic labels; Phase C maps evidence gaps and returns a ranked validation queue (feedback arrow); Phase D is the mandatory validation gate, drawn as a decision point: records that pass form the validated corpus with a quantitative screening error report; records that fail are excluded only with a documented reason family. MAPEVA-ScR inserts the machine phases *before* the human paradigm of the JBI scoping review methodology and wraps the human phase as a quantitative gate; it does not replace double human screening where resources permit (40) (39).

**Figure 1.**
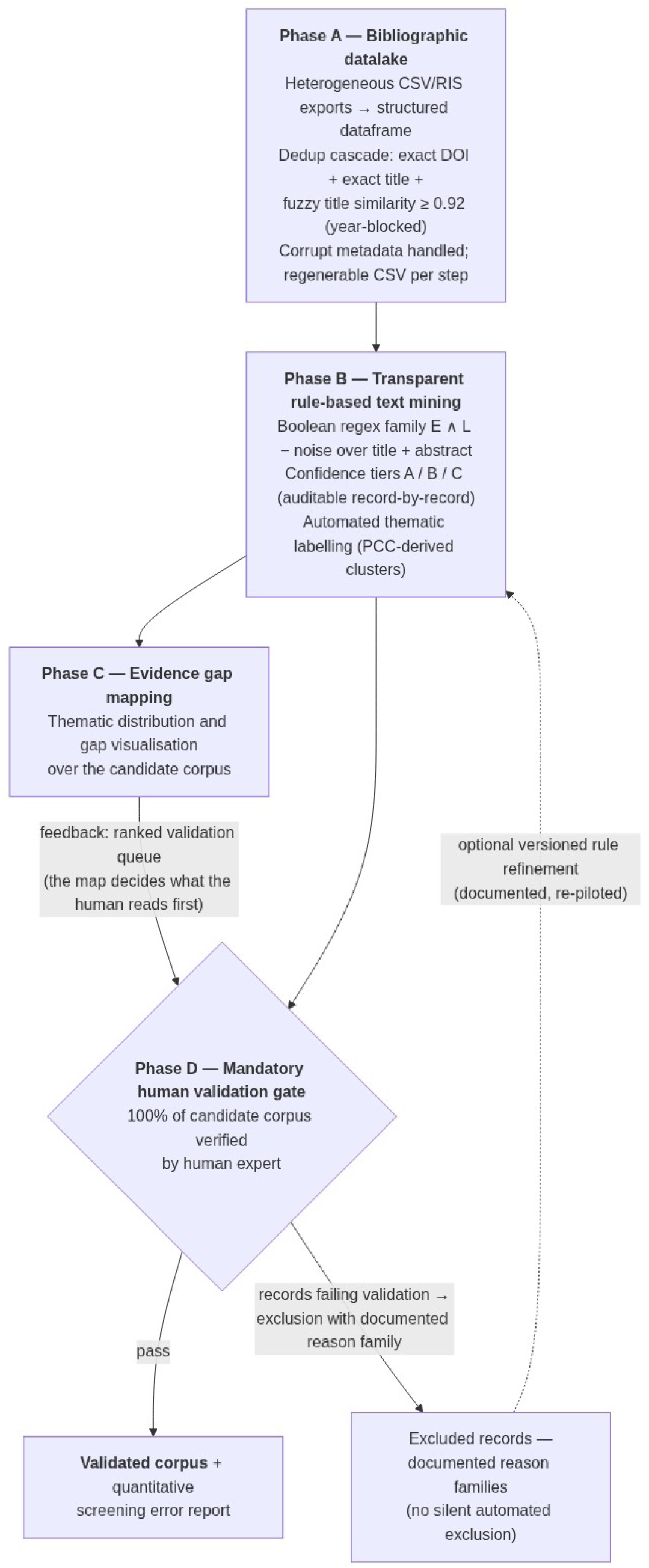
**The MAPEVA-ScR four-phase pipeline with the mandatory human validation gate.**The MAPEVA-ScR framework: four phases (A–D) with the feedback loop from Phase C mapping to human prioritisation and the mandatory validation gate (Phase D) as a decision point. The 0.92 fuzzy-deduplication threshold was chosen heuristically after inspecting candidate duplicate pairs during deduplication; a formal threshold sensitivity analysis is identified as future work.

Two features of Figure 1 deserve emphasis. First, the arrow from Phase C to Phase D is not decorative: the map orders the human’s reading, so the cells dominating the unvalidated candidate corpus and records nearest the decision boundary — Tier B and doubly flagged records — are verified first. Second, the decision diamond is the only point at which a record may leave the corpus, and only with a documented reason family; the dashed return arrow represents optional, versioned rule refinement, which always triggers re-validation rather than retroactive silent changes. The machine proposes and the human disposes: this operationalises the oversight requirement of the 2025 joint position statement of Cochrane, Campbell, JBI and the Collaboration for Environmental Evidence as a measurable artefact, not a narrative assurance (1).

### 3.2 Phase A — Datalake construction

Phase A converts heterogeneous bibliographic exports (CSV and RIS) into a single structured dataframe with harmonised columns (record identifier, source database, title, authors, year, DOI, abstract). Retrieval is documented with per-database quotas and known losses: in the case study, PubMed contributed 214 records, Scopus 794 and Web of Science 1,000 of 1,007 hits — seven records lost to the platform’s export limit, a marginal documented loss (present study). Search reporting follows PRISMA-S (38). Corrupt metadata (parsing artefacts, encoding errors, missing fields) are detected, corrected where possible, and otherwise flagged — never silently dropped.

Deduplication follows a deterministic cascade of increasing cost: (i) exact DOI match; (ii) exact match on normalised title (lowercased, punctuation and diacritics stripped); (iii) fuzzy title similarity ≥ 0.92 computed with Python’s `difflib.SequenceMatcher`, candidate pairs blocked by publication year to bound the pairwise comparisons; all fuzzy pairs are manually verified. The 0.92 threshold was chosen heuristically after inspecting candidate duplicate pairs during deduplication; a formal threshold sensitivity analysis is identified as future work. We chose transparent string similarity over probabilistic record linkage so that every merge decision remains reconstructible by a non-programmer reviewer. In the case study the cascade reduced 2,008 records to 1,435 uniques (−573 duplicates); a cross-check against Rayyan (1,005 candidate duplicates) was reconciled, the difference reflecting partial exports (present study). A date-window filter restricted the datalake to 2015–2025 — a ten-year recency window capturing contemporary occupational-health evidence, extending the period covered by the most recent systematic review of employability in epilepsy (50) — yielding 1,114 records (321 excluded by date; present study). Every step writes a regenerable CSV, so all downstream figures are traceable to source files.

### 3.3 Phase B — Transparent rule-based text mining with confidence tiers

#### 3.3.1 Rule design

The screening engine is a Boolean rule family of the form *E ∧ L − noise* applied as case-insensitive regular expressions over title and abstract: an entity axis *E* (terms for the health condition), a labour-context axis *L* (employment, fitness for work, workplace terms), and a noise set of known homonymy traps identified during iterative refinement. Rules were built from domain expertise and refined iteratively by inspecting the false positives detected during deduplication and pre-screening, with each refinement documented in version-controlled rule files; no formal pilot sample with inter-rater statistics was conducted, so this refinement is declared honestly as a partial analogue of the JBI piloting of eligibility criteria (40). Each rule application writes an auditable record (record identifier, matched pattern, rule version), so any inclusion can be traced to the exact substring that triggered it — a deliberate contrast with grey-box active learning (18), and closest in spirit to SWIFT-Review’s explicit topic filters (16), which we extend with tier stratification and the downstream gate.

#### 3.3.2 Tier stratification logic

Each retained record is assigned to a confidence tier: Tier A carries a direct signal (both axes matched strongly, typically in the title); Tier B a partial signal (one or both axes matched only in the abstract); Tier C a tangential one (a match via a broad contextual term, retained deliberately to protect sensitivity). Stratification serves only prioritisation of human reading and per-tier error measurement — never automatic exclusion, in line with the Cochrane caution (4). The logic is fully specified below.

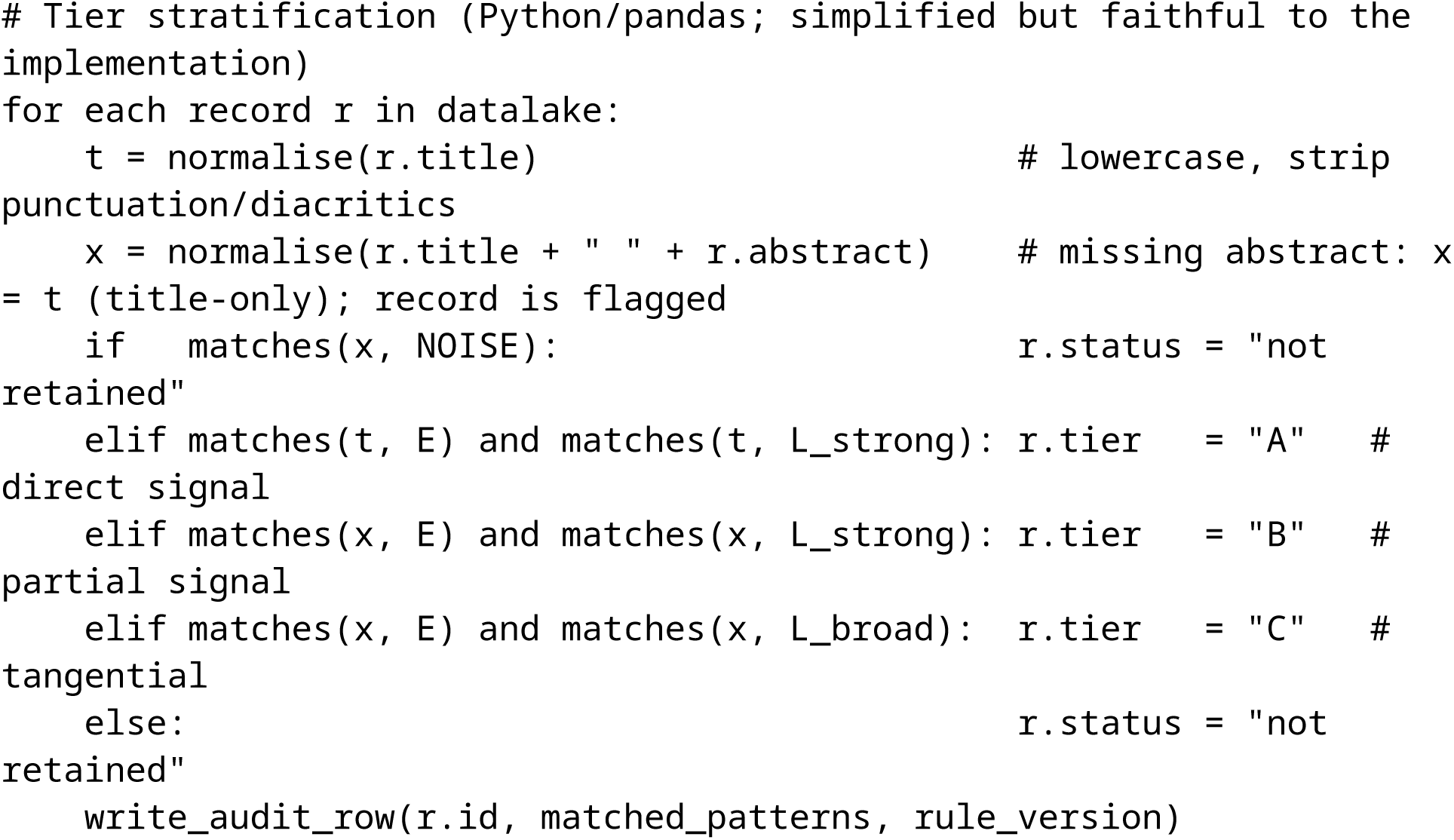

In the case study the filter retained 230 of 1,114 records (Tier A n = 102, Tier B n = 104, Tier C n = 24; present study). Records without an abstract were screened on the title alone and flagged as title-only. Non-retained records remain in the datalake, auditable and inspectable by the human verifier on demand.

#### 3.3.3 Automated thematic labelling

To feed the Phase C map, each candidate record receives provisional thematic labels from keyword clusters derived a priori from the PCC (Population–Concept–Context) question; Table 1 lists the four case-study clusters with example terms. An explicit warning is part of the framework: automated labels are hypotheses for the human gate, not results. In the case study the keyword set systematically under-labelled the Aptitude/employability cluster — an error visible only because the gate measured it (present study; §4.4).

**Table 1.**
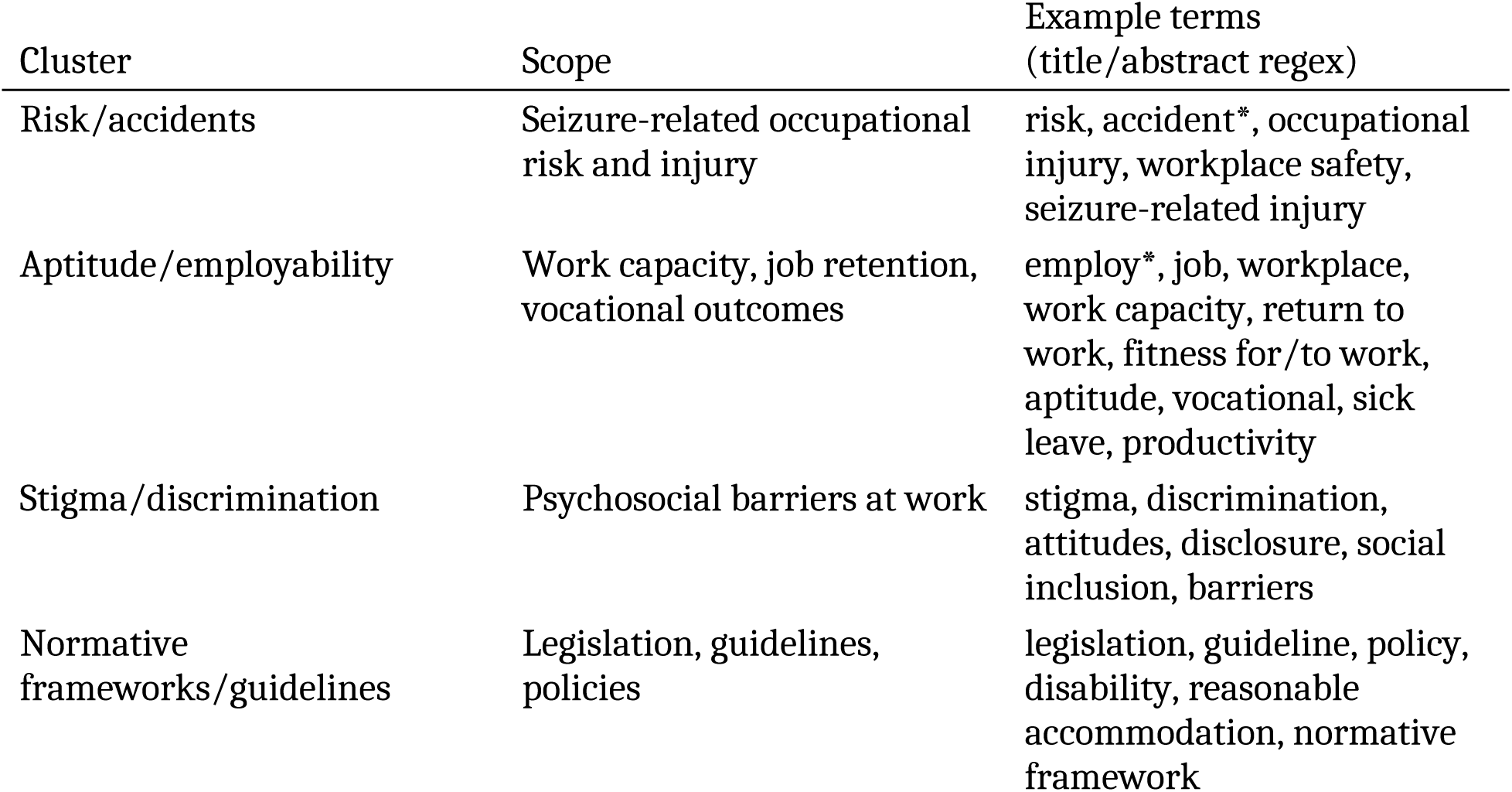
The four PCC-derived thematic keyword clusters for automated labelling, with example terms (case study implementation).

| Cluster | Scope | Example terms<br>(title/abstract regex) |
| --- | --- | --- |
| Risk/accidents | Seizure-related occupational risk and injury | risk, accident*, occupational injury, workplace safety, seizure-related injury |
| Aptitude/employability | Work capacity, job retention, vocational outcomes | employ*, job, workplace, work capacity, return to work, fitness for/to work, aptitude, vocational, sick leave, productivity |
| Stigma/discrimination | Psychosocial barriers at work | stigma, discrimination, attitudes, disclosure, social inclusion, barriers |
| Normative frameworks/guidelines | Legislation, guidelines, policies | legislation, guideline, policy, disability, reasonable accommodation, normative framework |

Three properties of the cluster design are worth noting. First, clusters are *a priori* and theory-driven (PCC), not emergent from topic modelling, so the map’s axes are interpretable before any data are seen and can be pre-registered. Second, labelling is multi-label: a record may carry several cluster tags, so cluster percentages need not sum to 100% — a property that must be stated in any map legend. Third, the terms are plain regex patterns in version-controlled text files, so a second reviewer can audit any label assignment; disagreements reduce to inspectable strings rather than model weights. The cost of this transparency is recall asymmetry across clusters — compact vocabularies (e.g., stigma terms) outperform diffuse ones (e.g., employability) — which is why labels are provisional and why the Phase D report includes a log of human label corrections.

### 3.4 Phase C — Evidence gap mapping with feedback

Phase C computes the thematic distribution of the candidate corpus across the clusters of Table 1 and visualises evidence density and gaps. The defining feature of MAPEVA-ScR is that this map *feeds back* into screening rather than being produced after it: it generates a ranked validation queue in which the human verifies first the cells that dominate the map (where errors would most distort conclusions) and records nearest the decision boundary (Tier B and doubly flagged records, where rule uncertainty concentrates). This contrasts with prevailing evidence gap map practice, which decouples mapping from screening — EPPI-Mapper maps after screening without feeding back prioritisation (20), and only 33% of Campbell evidence gap maps methodise gap identification at all (35). Phase C outputs two artefacts: the provisional map and the ranked validation queue consumed by Phase D.

The provisional character of the map is essential. In the case study, the pre-validation map ranked Risk/accidents first (19.1%), Normative frameworks/guidelines second (18.7%) and Aptitude/employability third (12.6%) — an ordering that human validation subsequently reversed (present study; full before/after figures in §4.4). Phase C output accordingly carries an explicit caveat: distributions are computed over the *unvalidated* candidate corpus and may not be reported as findings. The map directs verification effort to where its epistemic yield is highest, and the comparison between provisional and post-validation maps is itself a mandatory report item (item iv, below).

### 3.5 Phase D — Mandatory human validation gate and the screening error report

#### 3.5.1 The gate

Phase D requires human expert verification of 100% of the candidate corpus before any analytic use. Doubly flagged or doubtful records are resolved by documented expert judgement: in the case study, two doubtful records were resolved by the verifying specialist with written rationale (present study). Consistent with principle P2, no automated exclusion occurs without human sight of the candidate corpus. The gate operationalises the oversight requirement of the 2025 joint position statement (1) and the Cochrane Rapid Reviews Methods Group’s framing of AI as a quality-assurance adjunct (2), converting both from narrative assurances into a quantified, reportable procedure.

#### 3.5.2 The screening error report (proposed minimum reporting standard)

Passing the gate obliges the team to produce a screening error report with five mandatory contents: **(i)** a confusion matrix of automated tier × human judgement, with per-tier and global precision; **(ii)** a false-positive family taxonomy with counts, describing error structurally (e.g., homonymy families) rather than as a single aggregate; **(iii)** a log of human corrections to automated thematic labels; **(iv)** an explicit statement of which conclusions, maps or orderings changed after validation; and **(v)** traceability of every figure to regenerable CSVs and versioned rule files. Items (i)–(iv) are precisely what no current guideline requires (§2.3): PRISMA 2020 asks only that automation use be reported (12), and RAISE lists performance metrics as good practice for tool developers without mandating their report per synthesis (11). We propose the screening error report as a candidate minimum reporting standard for scoping reviews that use any automated screening component.

#### 3.5.3 Scalability rule

We state the scalability limit honestly. Full-corpus gating is feasible at the case study’s scale (230 candidate records, verified by a single domain expert within a normal screening workload). At approximately 5,000 candidates, 100% verification ceases to be proportionate, and the gate should be applied to a tier-stratified sample with extrapolated per-tier error bounds, preserving the report structure of §3.5.2. This sampled-gate variant is specified as future work (§5.3–5.4) and is deliberately *not* claimed as validated here: the present study provides empirical evidence only for full-corpus gating.

### 3.6 Appendix: screening rule sets (verbatim)

For full reproducibility, the exact rule sets used in the case study are reproduced verbatim below; they are plain regular expressions in version-controlled text files, applied as described in §3.3.

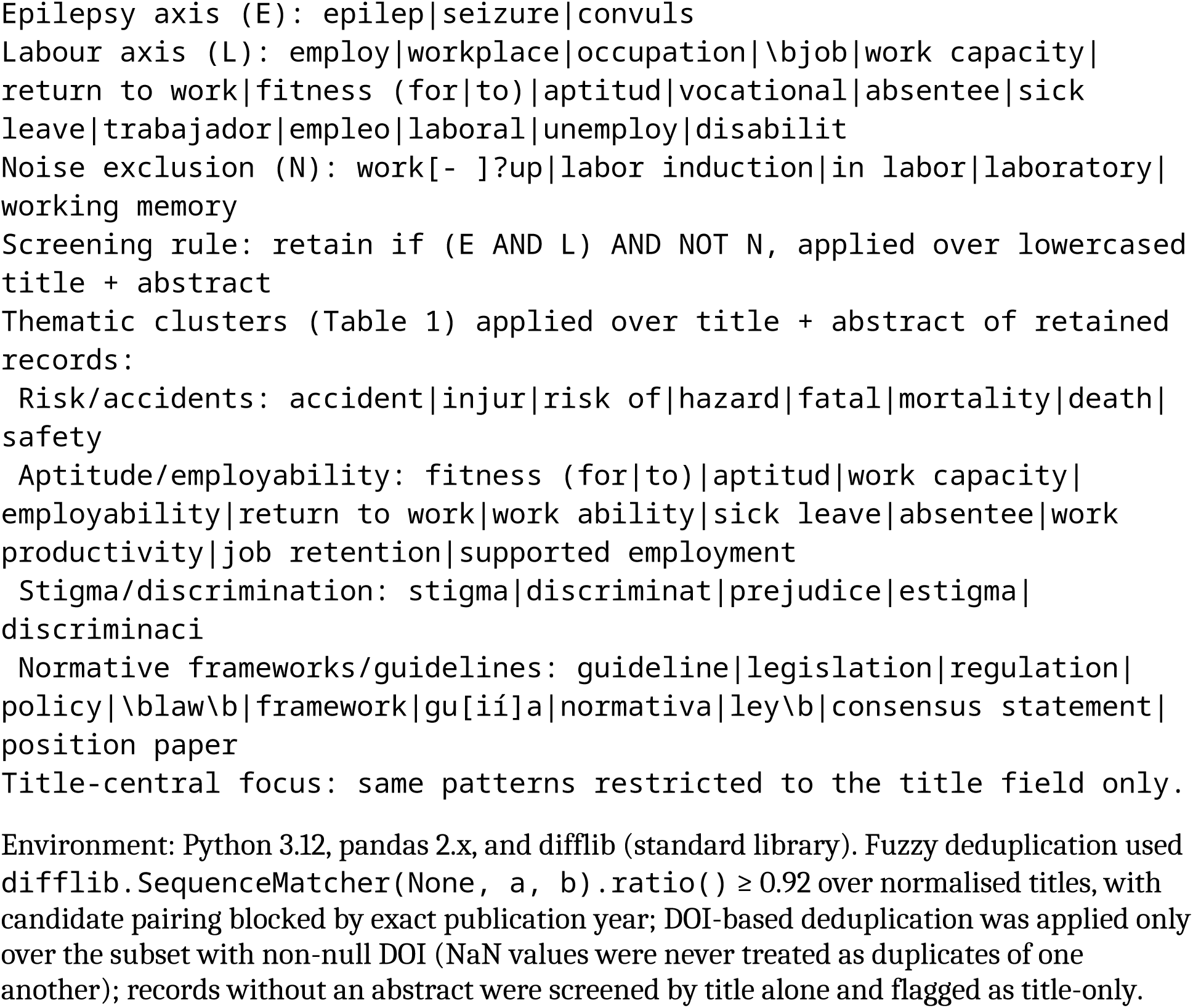

Environment: Python 3.12, pandas 2.x, and difflib (standard library). Fuzzy deduplication used `difflib.SequenceMatcher(None, a, b).ratio()` ≥ 0.92 over normalised titles, with candidate pairing blocked by exact publication year; DOI-based deduplication was applied only over the subset with non-null DOI (NaN values were never treated as duplicates of one another); records without an abstract were screened by title alone and flagged as title-only.

## 4. Empirical validation: case study — epilepsy and occupational fitness, 2015– 2025

### 4.1 Case question and protocol

The case study is a scoping review addressing the question: what is the state of the evidence on occupational fitness and employability of people with epilepsy or seizure disorders? Following the Population–Concept–Context (PCC) framework, the population comprised people with epilepsy or seizure disorders; the concept was occupational fitness, aptitude for work and employability; and the context was labour participation in any setting, restricted to publications from 2015–2025 retrieved from PubMed, Scopus and Web of Science. The domain was chosen deliberately: employability in epilepsy had previously been synthesised in a systematic review (50), providing a mature thematic literature against which an automated pipeline could err in informative ways, and no prior automated-methodological scoping review existed in this domain (present study). The human verifier was an occupational health specialist (the lead author). The data freeze dates were 7–8 September 2026; every figure reported below is regenerable from the source files Cribado_Epilepsia_PRECRIBADO.csv (n = 1,114) and Validacion_corpus_230_epilepsia.csv (n = 230) together with the evaluator’s human decision file.

### 4.2 Funnel and datalake

Figure 2 presents the case funnel. Multi-database retrieval yielded 2,008 records (PubMed 214, Scopus 794, Web of Science 1,000 of 1,007 hits; seven WoS records were not exported owing to the platform’s 1,000-record export limit — a marginal, documented loss). The deterministic deduplication cascade (exact DOI, exact normalised title, fuzzy title similarity ≥ 0.92; §3.2) reduced the datalake to 1,435 unique records; a cross-check against Rayyan, which flagged 1,005 candidate duplicates, was reconciled, the difference being explained by partial exports. The 2015–2025 date window excluded 321 records (Web of Science had been exported without a year filter), leaving 1,114 records. The rule filter (E ∧ L − noise; §3.3) retained 230 records as the candidate corpus, and the Phase D gate — expert verification of 100% of that corpus — confirmed 127. Because the human saw every candidate record, human judgement constitutes the reference standard and the machine’s contribution over the candidate corpus is measurable entirely as precision; the sensitivity of the upstream rule filter over the non-retained records is not measurable without human review of those records, and is addressed as a limitation (§5.3) (present study).

**Figure 2.**
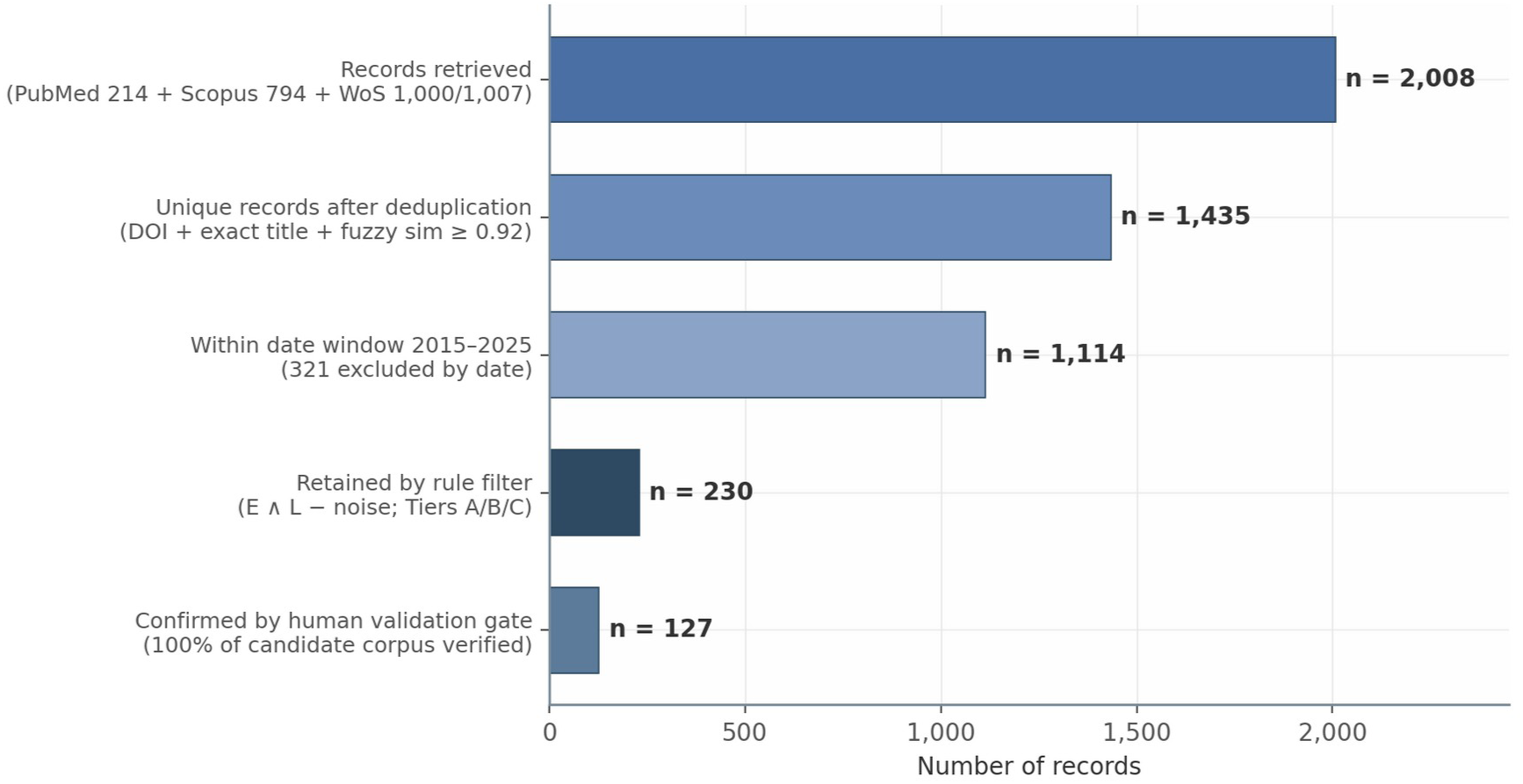
Screening funnel of the case study. Case-study screening funnel, epilepsy and occupational fitness, 2015–2025 (present study). Bars show the number of records surviving each successive stage, from multi-database retrieval to the human-validated corpus.

The funnel quantifies the division of labour that MAPEVA-ScR proposes. The machine stages reduced the corpus by 88.5% (2,008 → 230) before any human relevance decision was required; the gate then removed 103 spurious records from that reduced set. Two documented imperfections are deliberately retained rather than smoothed over: the seven unexported Web of Science records and the Rayyan discrepancy. Both are reported because a reproducible funnel must account for every record lost, not only those lost by design. All stage counts are regenerable from the source CSVs, satisfying the traceability item (v) of the screening error report (§3.5.2).

### 4.3 Screening error: confusion matrix and false-positive families

#### 4.3.1 Per-tier precision

Table 2 reports the cross-tabulation of automated tier assignment against human validation for the 230 candidate records.

**Table 2.** Cross-tabulation of automated tier assignment × human validation (n = 230; present study).

| Tier | Validated YES | Excluded NO | Total | Precision |
| --- | --- | --- | --- | --- |
| A (high confidence) | 77 | 25 | 102 | **75.5%** |
| B (medium confidence) | 39 | 65 | 104 | 37.5% |
| C (low confidence) | 11 | 13 | 24 | 45.8% |
| **Total** | **127** | **103** | **230** | **55.2%** |

*Note.* The false-negative quadrant is not estimable by design: non-retained records were not human-reviewed.

Three readings follow from Table 2. First, stratification orders risk as designed: Tier A precision (75.5%) is double that of Tier B (37.5%), confirming that the tier signal carries real information about likely relevance and can legitimately schedule the human’s verification queue. Second, even the highest tier carried 24.5% false positives, so “high confidence” is not publishable purity: without the gate, 103 spurious records — 44.8% of the candidate corpus — would have entered the analysis. Third, Tier C’s intermediate precision (45.8%) rests on only 24 records and should not be over-interpreted; its wide uncertainty is itself an argument for verifying rather than discarding the tangential tier. The practical consequence is that per-tier precision, not a single global figure, is the informative reporting unit.

#### 4.3.2 False-positive families and why the rules failed

Table 3 classifies the 103 false positives — records detectable as irrelevant only by the human verifier — into seven reproducible families.

**Table 3.** False-positive families identified by human validation (n = 103; present study).

| False-positive family | n | % |
| --- | --- | --- |
| Homonymy: ‘occupation’ in the military/territorial sense | 21 | 20.4% |
| Homonymy: ‘seizure’ as algorithmic/technical event detection (EEG, deep learning, robotics) | 16 | 15.5% |
| Homonymy: ‘seizure’ as legal confiscation | 14 | 13.6% |
| Clinical occupational therapy without a labour-market focus | 9 | 8.7% |
| Homonymy: ‘labor’ denoting childbirth (obstetrics) | 6 | 5.8% |
| Animal or in vitro models | 6 | 5.8% |
| Clinical or other records without a labour dimension | 31 | 30.1% |

The taxonomy exposes the mechanisms of rule failure. Four families are pure lexical polysemy that no keyword list can anticipate exhaustively: *occupation* in the military and territorial sense (21 records, 20.4%), *seizure* as the algorithmic detection of electroencephalographic events in the engineering literature (16, 15.5%), *seizure* as legal confiscation (14, 13.6%), and *labor* denoting childbirth (6, 5.8%). A fifth mechanism is semantic adjacency rather than homonymy: clinical occupational therapy shares its core vocabulary with occupational health while lacking any labour-market focus (9, 8.7%), and animal or in vitro models (6, 5.8%) match the entity axis perfectly while being structurally incapable of addressing the concept. The residual family — clinical records without a labour dimension (31, 30.1%) — reflects the inherent breadth of the labour axis. These families are domain-general and therefore predictable: they are precisely what a single aggregate black-box metric conceals and a family taxonomy exposes, and each maps to a specific, versioned amendment of the noise rule set for future iterations.

#### 4.3.3 Comparability with reported automated-screening error

The observed global precision of 55.2% is unremarkable against the published record. GPT-3.5 configured for sensitive screening reached specificity as low as 2.2% (5), and the Cochrane Rapid Reviews Methods Group documents incorrect AI inclusion decisions ranging from 0% to 29% (median 10%) across evaluations (2). Our case is therefore not an outlier; it is closer to the unreported norm of automated screening. This reading aligns with the long-standing caution of O’Mara-Eves and colleagues that text-mining gains in workload are purchased with sensitivity and precision costs that must be made explicit (3), and with the complementarity documented by Gates and colleagues, in which machine and human reviewers catch records the other misses (7). The difference in the present study is not the magnitude of the error but its measurement: the error was quantified against a full human gold standard and reported per tier and per family.

### 4.4 The reversed finding — before and after validation

#### 4.4.1 The inversion

Table 4 compares the thematic distribution computed over the unvalidated candidate corpus (n = 230) with that of the human-validated corpus (n = 127); labelling is multi-label, so percentages need not sum to 100%.

**Table 4.**
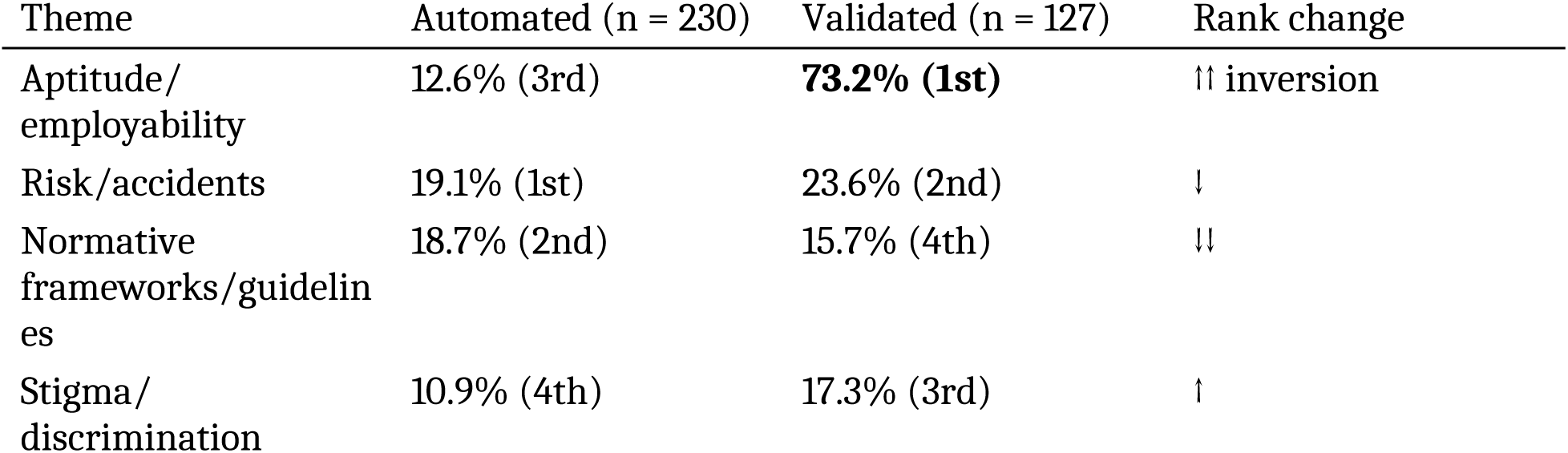
Thematic distribution before and after human validation, with rank changes (present study).

| Theme | Automated (n = 230) | Validated (n = 127) | Rank change |
| --- | --- | --- | --- |
| Aptitude/<br>employability | 12.6% (3rd) | **73.2% (1st)** | ↑↑ inversion |
| Risk/accidents | 19.1% (1st) | 23.6% (2nd) | ↓ |
| Normative<br>frameworks/guidelines | 18.7% (2nd) | 15.7% (4th) | ↓↓ |
| Stigma/<br>discrimination | 10.9% (4th) | 17.3% (3rd) | ↑ |

Table 4 contains the central empirical argument of this article. The unvalidated screening would have supported the conclusion that “the literature prioritises risk over aptitude”: Risk/accidents ranked first and Aptitude/employability only third. After the gate, the validated conclusion is reversed in rank and different in substance: employability dominates in the broad sense (73.2% of included records), yet standardised aptitude criteria (11.8% of titles) and stigma (4.7% of titles) as central, title-level foci remain marginal. Both statements are quantitative rather than narrative: the false conclusion is attributable to a measured error (an under-labelled cluster plus 44.8% contaminating false positives), not to an unspecified “automation problem”. A team reporting the pre-validation map would have published a thematically inverted synthesis while remaining nominally compliant with current reporting expectations.

#### 4.4.2 Why the inversion happened

The inversion has two additive sources, both measured. First, the automated keyword set systematically under-labelled Aptitude/employability: 107 of the 127 included records required human correction of their thematic labels (present study). The mechanism is recall asymmetry across clusters (§3.3.3): risk and regulatory vocabularies are compact (“risk”, “accident”, “legislation”, “guideline”), whereas employability is expressed diffusely — job retention, work capacity, vocational rehabilitation, return to work, sick leave, reasonable adjustment — so a fixed keyword set captures risk language far more completely than aptitude language. Second, the 103 false positives were not thematically neutral: homonymous and clinically adjacent records inflated the apparent density of the non-aptitude cells of the map. Removing them while correcting labels moved Aptitude/employability from 12.6% to 73.2% — a shift of more than sixty percentage points produced entirely by validation, with no change to the underlying literature.

#### 4.4.3 Human correction work and traceability

The gate’s workload is itself data. Beyond the 107 label corrections, two doubly flagged “doubtful” records were resolved by the verifying specialist’s documented clinical judgement, and a double independent control — human decision plus automated check — confirmed that 0 of the 127 included records lacks an epilepsy term in title or abstract. One discrepancy was detected and handled transparently: the evaluator self-reported 110 validated records against 127 in the decision file; the discrepancy was documented and resolved by adopting the primary source (the file). We report these frictions deliberately. A validation gate that is described only through its clean outputs would reproduce, at the human stage, the same opacity the framework condemns at the machine stage.

### 4.5 Effort and feasibility

The full-corpus gate on 230 records was feasible for a single domain expert within a normal screening workload, and the gate itself entails no sensitivity loss over the candidate corpus — the human saw 100% of it. The sensitivity of the upstream rule filter over the 884 non-retained records is unknown by design and is addressed as a limitation (§5.3). This stands in contrast to the ∼5% sensitivity loss tolerated as a known cost of semi-automation in the Cochrane Handbook (4). We state the feasibility limits honestly: a single expert verifier performed the gate, so no inter-rater reliability coefficient could be computed; this was mitigated, but not eliminated, by the automated double control reported above, and is taken up as a limitation in §5.3. Scalability beyond a few hundred candidates requires the tier-stratified sampled gate specified in §3.5.3, for which the per-tier precision estimates of Table 2 provide exactly the inputs needed to compute extrapolated error bounds.

## 5. Discussion

### 5.1 Principal findings and interpretation

Three findings emerge from the empirical validation, each with a distinct implication for evidence synthesis practice. First, transparent rule-based screening achieved useful prioritisation but not publishable purity: the tier signal ordered risk as designed (Tier A precision 75.5% against 37.5% in Tier B), yet even the highest-confidence tier carried 24.5% false positives, and the candidate corpus as a whole contained 103 spurious records — 44.8% contamination that only the human gate removed (present study). Rules of the *E ∧ L − noise* family are therefore best understood as a workload-reduction and scheduling device whose output is a hypothesis, not a result. Second, the error was structured rather than random: the 103 false positives collapsed into seven reproducible families, dominated by domain-general lexical homonymy (‘occupation’ in the military sense, ‘seizure’ as algorithmic event detection, ‘labor’ as childbirth). Structured error is reportable and correctable — each family maps to a versioned amendment of the noise set — whereas a single aggregate accuracy figure would have concealed exactly the information needed to improve the filter. Third, and most consequentially, the error did not remain at the screening stage: it propagated to a reversed thematic conclusion, with Aptitude/employability moving from 12.6% (third) to 73.2% (first) after validation (present study). This is a consequence-level harm — a published synthesis with an inverted substantive claim — not merely a workload or efficiency concern.

These findings add a consequence-level reading to the comparability analysis of §4.3.3. The sensitivity and precision costs that O’Mara-Eves and colleagues warned must be made explicit rather than assumed (3) materialised here not as workload or efficiency losses but as a conclusion-level harm — a published synthesis with an inverted substantive claim. The defensible inference is not that automated screening is unusually poor, but that its error is large enough to invert conclusions and is almost never measured where it is used.

### 5.2 Relation to guidelines and honesty nuances

Two honesty obligations frame the contribution. First, human oversight is not our invention: the 2025 joint position statement of Cochrane, the Campbell Collaboration, JBI and the Collaboration for Environmental Evidence already mandates it (1). What MAPEVA-ScR contributes is the conversion of that narrative requirement into a mandatory validation gate with quantitative error metrics — per-tier precision, a confusion matrix against a human gold standard, a false-positive family taxonomy, and an explicit statement of which conclusions changed after validation. In this sense the framework aligns with, and operationalises, the governance trend rather than contesting it. Second, the regulatory gap must be formulated precisely: it is the absence of a mandatory, quantitative reporting requirement *as of today*, not an eternal void. RAISE 2 already lists performance metrics as good practice for tool developers and evaluators (11), PRISMA-trAIce has been proposed as a dedicated reporting checklist (14), and the Cochrane Rapid Reviews Methods Group requires an AI Use Disclosure while positioning AI as quality assurance (2). The gap is closing; MAPEVA-ScR offers an operational template that review teams and journals can adopt immediately, without waiting for the convergence of these initiatives.

We also stress complementarity with the active-learning paradigm rather than competition. Nothing in MAPEVA-ScR precludes placing the validation gate after an ASReview-style prioritisation (18): the gate measures the error of whatever upstream component produced the candidate corpus, whether rule-based, classifier-based or LLM-based. The screening error report (§3.5.2) is thus upstream-agnostic and could serve as a minimum reporting standard for any scoping review that uses an automated screening component — which is precisely the property a guideline extension requires.

### 5.3 Limitations

Several limitations qualify these claims. The empirical evidence rests on a single case study in a single domain (epilepsy × occupational health); the magnitude of the reversal, though not the existence of error, may not generalise to topics with more uniform vocabularies. The gate was executed by a single expert verifier, so no inter-rater reliability coefficient (e.g., Cohen’s kappa) could be computed; this was mitigated, but not eliminated, by the automated double control confirming that all 127 included records contained an epilepsy term. The rule-based design deliberately trades precision for transparency: a trained classifier might exceed 55.2% precision, but at the cost of record-level auditability, which we judge the more valuable property for a validation-centred framework. The corpus was moderate (230 candidates): full-corpus gating is feasible at this scale, but beyond roughly 5,000 candidates a 100% human gate ceases to be proportionate, and the tier-stratified sampled gate with extrapolated error bounds sketched in §3.5.3 remains untested future work — we do not claim validation for that variant. The sensitivity of the rule-based filter was not measured; a human review of a stratified sample of the 884 non-retained records is required to bound recall, and is planned as the next validation step. Rules are domain-built and require re-piloting for each new topic, mirroring the JBI piloting logic (40). Our positioning search was broad but not formally systematic, so the novelty claim is phrased as “to our knowledge” pending verification in Scopus and Web of Science, and the acronym check for MAPEVA-ScR likewise awaits formal confirmation. Finally, the funnel itself contains documented imperfections — seven unexported Web of Science records and an evaluator self-report discrepancy (110 versus 127) resolved from the primary file — which we report rather than smooth over.

### 5.4 Implications and future work

Three implications follow. For non-programmer occupational and public health researchers, MAPEVA-ScR demonstrates that an auditable automation pipeline is within reach of a Python/pandas skill set, and that the human expertise they already possess is the component that converts screening output into defensible evidence. For editors and guideline developers, the screening error report is a concrete candidate for a PRISMA-ScR or PRISMA-AI extension item; journals could plausibly require it as a submission condition for reviews reporting automated screening, just as flow diagrams are required today (12). For the field, the timing argument is urgent: oversight is already mandated, quantitative reporting is being actively designed (11) (14), and the window in which unmeasured screening error can pass unremarked is closing — teams that instrument their pipelines now will be compliant with the standards that are coming, not merely those that exist. Future work comprises formal multi-case replication across domains, dual-verifier designs enabling inter-rater reliability estimation, validation of the tier-stratified sampled gate at corpora of ∼5,000 records, and integration studies pairing the gate with active-learning prioritisation and with living and rapid review formats (2).

## 6. Conclusions

MAPEVA-ScR converts human oversight — already mandated by the 2025 joint statement of Cochrane, Campbell, JBI and the Collaboration for Environmental Evidence but nowhere required to be measured (1) — into a mandatory validation gate with quantitative error reporting. The empirical case shows why this matters. A transparent rule-based filter reduced 2,008 records to a candidate set of 230, but at a global precision of 55.2% (Tier A 75.5%, B 37.5%, C 45.8%), leaving 103 false positives (44.8%) across seven reproducible error families (present study). Without the gate, the thematic conclusion would have been published reversed: automated labelling ranked Aptitude/employability third (12.6%) when human validation placed it first (73.2%), after 107 of 127 included records required label correction (present study). The full pipeline — datalake, tier-stratified rule-based text mining, feedback gap mapping, and the gated error report — is executable by non-programmer researchers, with every decision auditable record by record.

The takeaway is a single sentence: the machine proposes, the human disposes — and the field must require proof that the human disposed. We therefore propose the screening error report as a minimum reporting standard for any evidence synthesis using automated screening, filling a gap that PRISMA 2020, PRISMA-ScR, JBI and MECIR leave open as of today.

Future work comprises four directions: multi-validator replication with inter-rater reliability coefficients; tier-stratified sampled gating with extrapolated error bounds for larger corpora; formal Scopus and Web of Science verification of the novelty claim before submission; and a prospective adoption study of the gate’s feasibility and cost across teams and domains.

## Declarations

## Ethics approval and consent to participate

Not applicable; this is a methodological study conducted exclusively on published literature.

## Consent for publication

Not applicable.

## Availability of data and materials

The screening datasets (Cribado_Epilepsia_PRECRIBADO.csv, n = 1,114; Validacion_corpus_230_epilepsia.csv, n = 230), validation logs and Python/pandas code are available from the corresponding author on request and will be deposited in an open repository [OSF/Zenodo DOI — TO COMPLETE] before publication.

## Authors’ contributions

CRediT. César Jesús Eras Lévano: conceptualisation, methodology, data curation, human verification, writing — original draft. Remaining roles to be assigned.]

## Data Availability

All data produced in the present work are contained in the manuscript

## Acknowledgements

None

## AI Use Disclosure

Generative AI tools assisted with language editing and drafting of portions of this manuscript under full human verification, in line with the Cochrane Rapid Reviews Methods Group position on AI use disclosure (2); all quantitative data were verified by the authors against the primary source files. The MAPEVA-ScR pipeline itself uses deterministic rule-based text mining; no generative AI participates in screening decisions.

